# Proteomic profiling of peripheral blood mononuclear cells reveals immune dysregulation and metabolic alterations in kidney transplant recipients with COVID-19

**DOI:** 10.1101/2024.09.19.24313795

**Authors:** Giuseppe G. F. Leite, Mônica Bragança Sousa, Larissa de Oliveira C. P. Rodrigues, Milena Karina Colo Brunialti, José Medina-Pestana, Joe M. Butler, Hessel Peters-Sengers, Lúcio Requião-Moura, Reinaldo Salomão

## Abstract

The emergence of the Coronavirus disease 2019 (COVID-19) pandemic in 2020 has profoundly impacted global health systems, particularly affecting vulnerable populations like kidney transplant recipients (KTRs).We prospectively collected blood samples from 17 PCR-confirmed COVID-19 KTR patients and 10 non-COVID-19 KTRs between May and September 2020. Using tandem mass tag-based quantitative proteomics, we characterized peripheral blood mononuclear cells (PBMCs) from KTRs alongside plasma protein biomarkers and lymphocyte counts, followed by bioinformatics analyses. Our study revealed significant proteomic alterations within PBMCs of SARS-CoV-2 infected KTRs, particularly in pathways associated with glycolysis, glucose metabolism, and neutrophil degranulation. Additionally, we observed an altered immune response marked by elevated cytokines and inflammatory mediators, coupled with decreased lymphocyte counts. Notably, patients with acute kidney injury (AKI) exhibited worse outcomes, including higher rates of ICU transfer and mechanical ventilation. Comparison of PBMC proteomic profiles between AKI and non-AKI patients highlighted distinct immune-related pathways, with AKI patients showing pronounced alterations in innate immune responses, particularly in neutrophil degranulation. Moreover, our analysis unveiled a negative correlation between T cell counts and neutrophil degranulation, suggesting potential implications for immune dysregulation in COVID-19. Our findings shed light on the complex proteomic landscape and immune responses in COVID-19-infected KTRs, emphasizing the critical need for studies focused on this population, especially in individuals with AKI. Furthermore, our observations provide valuable insights for further exploration of therapeutic interventions targeting immune dysregulation pathways in this vulnerable population.

## INTRODUCTION

The Coronavirus disease 2019 (COVID-19) pandemic, which emerged in 2020, rapidly became a global health crisis, profoundly affecting healthcare systems, economies, and overall quality of life [1, 2]. Prior to the achievement of widespread vaccination coverage, COVID-19 presented with a wide range of clinical manifestations, from asymptomatic or presymptomatic states to varying degrees of severity, including mild, moderate, severe, and, in some cases, life-threatening complications[3].

Individuals with compromised immune systems, especially those undergoing immunosuppressive therapy, were particularly vulnerable. Among these high-risk groups, kidney transplant recipients (KTRs) demonstrated notably higher case-fatality rates, even after vaccination [4, 5], and exhibited lower seroconversion rates following vaccination [6]. This elevated risk underscores the compounded vulnerabilities associated with immunosuppressive treatments and the presence of multiple comorbidities in these patients [7–9].

The cumulative number of comorbidities associated with an immune system impairment has led to diverse responses to COVID-19 among KTRs compared with non-transplanted patients [7–9]. Investigating a panel of circulating cytokines and vascular mediators in sequential samples, we previously demonstrated that both non-KTRs and KTRs infected with COVID-19 exhibited a similar transition from admission to convalescent samples. However, the key mediators differentiating patients from their respective controls (i.e., healthy volunteers and non-infected KTRs) were distinct. For non-KTRs, the top differentiators were inflammatory cytokines, whereas for KTRs, endothelial response patterns and levels of Neutrophil Gelatinase-Associated Lipocalin (NGAL)[10], an early marker of acute kidney injury (AKI), were the most significant. AKI, commonly observed in numerous systemic conditions [11], has been linked to poor outcomes, a trend that persisted in COVID-19 patients. A multicenter study involving 1,680 KTRs with COVID-19 revealed that the mortality rate was 36.0% in those with AKI, compared to 19.1% in patients maintaining normal renal function, highlighting renal function as a key predictor of survival in this population [12].

Recently, mass spectrometry (MS)-based proteomics, primarily focused on plasma, serum, and urine, has emerged as a powerful tool to shed light on a broad spectrum of dysregulated biological processes in KTRs with COVID-19 [13, 14]. However, the influence of SARS-CoV-2 infection on peripheral blood mononuclear cells (PBMCs) in KTRs remains a subject of ongoing investigation.

In this study, we employed a tandem mass tag (TMT)-based quantitative proteomic approach, coupled with bioinformatics analyses, to investigate altered proteins associated with dysregulated pathways and biological processes in kidney transplant recipients (KTRs) with COVID-19. We focused on patients with moderate to severe illness admitted to hospital wards who presented with or without acute kidney injury (AKI), as well as patients discharged from the hospital. Our aim was to provide insights into proteomic changes during the early stages of the disease and their association with AKI and post-clinical recovery.

## MATERIALS AND METHODS

### Study Design and Population

This prospective cohort study was conducted at Hospital São Paulo, a tertiary university hospital in São Paulo, Brazil [15]. The study was approved by the research ethics committee (Process number 4.453.137), and all volunteers provided written informed consent prior to enrolment. The study comprised adult patients (≥18 years old) diagnosed with COVID-19 based on positive reverse transcriptase polymerase chain reaction (RT-PCR) results from nasopharyngeal swabs. These patients were admitted to the hospital wards between May 10 and September 26, 2020. This period coincided with the first wave of the COVID-19 pandemic in Brazil (February 25, 2020, to November 5, 2020)[16], when the predominant variants observed were B.1.1.28 (20–30%) and B.1.1.33 (10–35%)[16]. Patients included in the cohort had moderate to severe illness, as defined by guidelines from the National Institutes of Health and the World Health Organization[17, 18]. Exclusion criteria encompassed patients referred to outpatient clinics or admitted directly to the ICU. Among the initial 68 COVID-19 patients in the cohort, 17 KTRs were selected for this study (for a comprehensive overview of the entire cohort, please refer to Peçanha-Pietrobom et al [15]). Additionally, a control group comprised 10 KTR volunteers without COVID-19 symptoms (KTR controls), matched to the patient group for time since transplantation, immunosuppressive therapy, sex, and age. None of the participants in this study were vaccinated due to the unavailability of vaccines at the time of enrolment.

AKI was prospectively defined according to the Kidney Disease Improving Global Outcomes (KDIGO) criteria [19, 20]. Baseline graft function was assessed using creatinine levels recorded three months prior to COVID-19 diagnosis, and AKI was identified based on creatinine levels at hospital admission.

### Sample Collection

Blood samples were collected from both patients and KTR controls into ethylenediaminetetraacetic acid (EDTA)-treated tubes (BD Biosciences, San Diego, CA, USA). Plasma and peripheral blood mononuclear cells (PBMCs) were separated using a Ficoll gradient method (Ficoll-Paque PLUS, GE Healthcare Biosciences, Uppsala, Sweden). Plasma samples were stored at –80°C, and PBMCs were preserved in liquid nitrogen for subsequent use.

### Measurements

For detailed information on the proteomics experiments, data processing protocols, plasma biomarker assays, and the corresponding analysis methods, please refer to the supplementary methods in **Supplementary Material 1.**

### Statistical Analysis

Statistical analyses were performed using R (version 4.3.0). Normality of data was assessed using the Shapiro–Wilk test and Quantile–Quantile (Q-Q) plots. Non-normally distributed data were analyzed using the Mann–Whitney U test or the Kruskal–Wallis test, while normally distributed data were analyzed using Welch’s t-test or analysis of variance (ANOVA). For post hoc testing of non-normally distributed data, Dunn’s test of multiple comparisons using rank sums was employed; for normally distributed continuous data, Tukey’s post hoc test was conducted. Categorical variables were compared using the chi-square test or Fisher’s exact test, as appropriate. Biomarker data were log-transformed to normalize their distribution. To visualize the overall differences among plasma biomarkers, principal component analysis (PCA) was conducted following previously established method [21, 22]. Correlation analyses were performed using Spearman’s rank correlation coefficient (Rho). Differences between groups were quantified using standardized mean differences (SMD) [23].

### Bioinformatics

Differential protein abundance analysis was performed using the R/Bioconductor package limma. The model was adjusted for additional covariates: age, sex, time after transplantation (months), maintenance immunosuppressive therapy (class), and donor status (deceased or living). Differences in PBMC proteome profiling between the two groups were detected using the empirical Bayes moderated t-statistics, and Benjamini-Hochberg corrections were applied to all p-values to calculate the false discovery rates (FDR). Protein set enrichment analysis (PSEA) was performed using the fgsea package, with annotation information from the Reactome pathway database and hallmark gene sets from MSigDB. Additionally, single-sample PSEA (ssPSEA) was performed using the hacksig package and the Reactome pathway database. We focused on Reactome-defined immune-related pathways implicated in the host response to infection: innate immune system, adaptive immune system, cytokine signaling in the immune system, hemostasis, programmed cell death, and metabolism.

## RESULTS

### Clinical and Epidemiological Features of the Study Participants

Seventeen kidney transplant recipients (KTRs) with COVID-19, a subset of a hospital-admitted cohort with diverse clinical outcomes[15] were analyzed in this study (**Table 1**). According to the KDIGO criteria for staging AKI, we observed that six patients (35%) did not present with AKI (stage 0), while five (29%) were classified as stage 1, three (17%) as stage 2, and three (17%) as stage 3. The mean age was 54.0 years, and most were male (n = 10, 59%). The time between COVID-19 symptom onset and hospital admission was 7.5 ± 4.6 days. Primary symptoms included dyspnea (n = 12, 71%), cough (n = 12, 71%), fever (n = 7, 41%), diarrhea (n = 7, 41%), and nausea/vomiting (n = 7, 41%). Arterial hypertension was the most prevalent comorbidity (n = 13, 76%). Additionally, 53% of patients required ICU admission, 41% underwent mechanical ventilation, 29% required renal replacement therapy, and 41% died. Among the survivors, six participated in a clinical follow-up at an average of 29.3 ± 5.0 days after hospital discharge (designated as Convalescent Sample 30 [CS30]). No significant differences were observed in demographic data, maintenance immunosuppressive therapy class, or donor status when comparing KTRs with COVID-19 and KTR controls (**Table 1**).

**Table 1.**
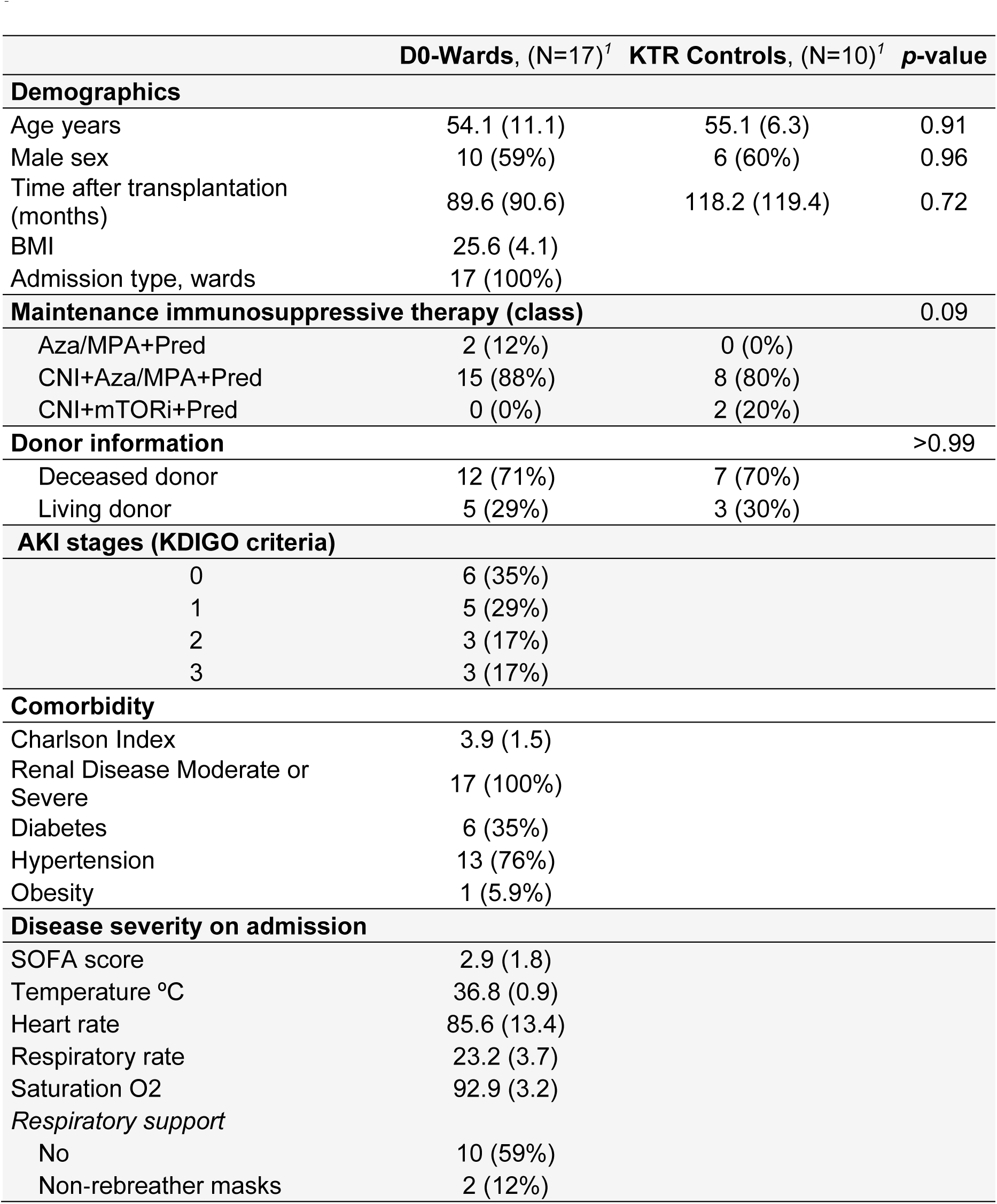
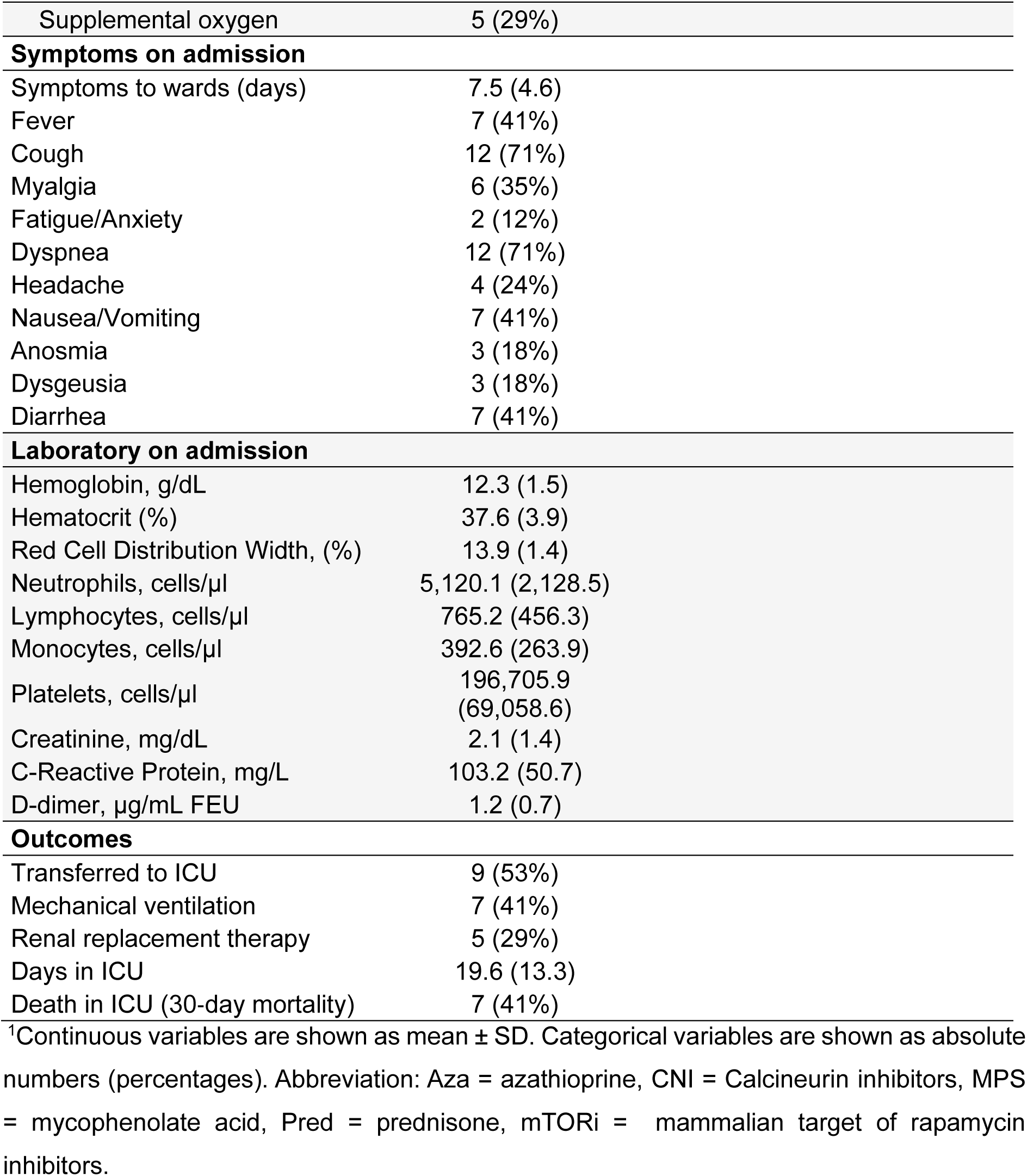
Baseline characteristics and clinical outcomes of KTR COVID-19 patients at ward admission and KTR Controls.

|  | D0-Wards, (N=17)<sup>1</sup> | KTR Controls, (N=10)<sup>1</sup> | p-value |
| --- | --- | --- | --- |
| **Demographics** |  |  |  |
| Age years | 54.1 (11.1) | 55.1 (6.3) | 0.91 |
| Male sex | 10 (59%) | 6 (60%) | 0.96 |
| Time after transplantation (months) | 89.6 (90.6) | 118.2 (119.4) | 0.72 |
| BMI | 25.6 (4.1) |  |  |
| Admission type, wards | 17 (100%) |  |  |
| **Maintenance immunosuppressive therapy (class)** |  |  | 0.09 |
| Aza/MPA+Pred | 2 (12%) | 0 (0%) |  |
| CNI+Aza/MPA+Pred | 15 (88%) | 8 (80%) |  |
| CNI+mTORi+Pred | 0 (0%) | 2 (20%) |  |
| **Donor information** |  |  | >0.99 |
| Deceased donor | 12 (71%) | 7 (70%) |  |
| Living donor | 5 (29%) | 3 (30%) |  |
| **AKI stages (KDIGO criteria)** |  |  |  |
| 0 | 6 (35%) |  |  |
| 1 | 5 (29%) |  |  |
| 2 | 3 (17%) |  |  |
| 3 | 3 (17%) |  |  |
| **Comorbidity** |  |  |  |
| Charlson Index | 3.9 (1.5) |  |  |
| Renal Disease Moderate or Severe | 17 (100%) |  |  |
| Diabetes | 6 (35%) |  |  |
| Hypertension | 13 (76%) |  |  |
| Obesity | 1 (5.9%) |  |  |
| **Disease severity on admission** |  |  |  |
| SOFA score | 2.9 (1.8) |  |  |
| Temperature °C | 36.8 (0.9) |  |  |
| Heart rate | 85.6 (13.4) |  |  |
| Respiratory rate | 23.2 (3.7) |  |  |
| Saturation O2 | 92.9 (3.2) |  |  |
| **Respiratory support** |  |  |  |
| No | 10 (59%) |  |  |
| Non-rebreather masks | 2 (12%) |  |  |

|  |  |
| --- | --- |
| Supplemental oxygen | 5 (29%) |
| **Symptoms on admission** |  |
| Symptoms to wards (days) | 7.5 (4.6) |
| Fever | 7 (41%) |
| Cough | 12 (71%) |
| Myalgia | 6 (35%) |
| Fatigue/Anxiety | 2 (12%) |
| Dyspnea | 12 (71%) |
| Headache | 4 (24%) |
| Nausea/Vomiting | 7 (41%) |
| Anosmia | 3 (18%) |
| Dysgeusia | 3 (18%) |
| Diarrhea | 7 (41%) |
| **Laboratory on admission** |  |
| Hemoglobin, g/dL | 12.3 (1.5) |
| Hematocrit (%) | 37.6 (3.9) |
| Red Cell Distribution Width, (%) | 13.9 (1.4) |
| Neutrophils, cells/μl | 5,120.1 (2,128.5) |
| Lymphocytes, cells/μl | 765.2 (456.3) |
| Monocytes, cells/μl | 392.6 (263.9) |
| Platelets, cells/μl | 196,705.9<br>(69,058.6) |
| Creatinine, mg/dL | 2.1 (1.4) |
| C-Reactive Protein, mg/L | 103.2 (50.7) |
| D-dimer, μg/mL FEU | 1.2 (0.7) |
| **Outcomes** |  |
| Transferred to ICU | 9 (53%) |
| Mechanical ventilation | 7 (41%) |
| Renal replacement therapy | 5 (29%) |
| Days in ICU | 19.6 (13.3) |
| Death in ICU (30-day mortality) | 7 (41%) |

<sup>1</sup>Continuous variables are shown as mean ± SD. Categorical variables are shown as absolute numbers (percentages). Abbreviation: Aza = azathioprine, CNI = Calcineurin inhibitors, MPS = mycophenolate acid, Pred = prednisone, mTORi = mammalian target of rapamycin inhibitors.

**Table 2.** Demography, comorbidities, clinical and laboratory admission data, and outcomes stratified according to the presence of acute kidney injury.

| Characteristic | AKI (N=11)<sup>†</sup> | non-AKI (N=6)<sup>†</sup> | SMD |
| --- | --- | --- | --- |
| **Demographics** |  |  |  |
| Age years | 51.7 (12.4) | 58.2 (7.4) | -0.56 |
| Male sex | 7 (64%) | 3 (50%) | 0.13 |
| BMI | 24.8 (4.4) | 26.9 (3.7) | -0.53 |
| **Comorbidity** |  |  |  |
| Charlson Index | 3.8 (1.7) | 4.0 (1.4) | -0.12 |
| Diabetes | 3 (27%) | 3 (50%) | -0.48 |
| Hypertension | 7 (64%) | 6 (100%) | -1.07 |
| Obesity | 0 (0%) | 1 (17%) | -0.63 |
| **Disease severity on admission** |  |  |  |
| SOFA score | 3.5 (1.8) | 2.0 (1.4) | 0.90 |
| Temperature °C | 36.9 (1.0) | 36.6 (0.6) | 0.42 |
| Heart rate | 83.8 (15.1) | 88.8 (9.9) | -0.39 |
| Respiratory rate | 22.9 (4.6) | 23.8 (1.3) | 0.27 |
| Saturation O2 | 92.8 (3.6) | 93.0 (2.5) | -0.06 |
| *Respiratory support* | 4 (36.4%) | 3 (50.0%) | 0.28 |
| **Symptoms on admission** |  |  |  |
| Symptoms to wards (days) | 8.1 (5.5) | 6.5 (2.1) | 0.38 |
| Fever | 5 (45%) | 2 (33%) | 0.25 |
| Cough | 7 (64%) | 5 (83%) | -0.46 |
| Myalgia | 5 (45%) | 1 (17%) | 0.66 |
| Fatigue/Anxiety | 1 (9.1%) | 1 (17%) | -0.23 |
| Dyspnea | 7 (64%) | 5 (83%) | 0.46 |
| Headache | 2 (18%) | 2 (33%) | 0.35 |
| Vomiting/nausea | 4 (36%) | 3 (50%) | 0.28 |
| Anosmia | 1 (9.1%) | 2 (33%) | 0.62 |
| Dysgeusia | 1 (9.1%) | 2 (33%) | 0.62 |
| Diarrhea | 4 (36%) | 3 (50%) | 0.28 |
| **Laboratory on admission** |  |  |  |
| Hemoglobin, g/dL | 12.1 (1.5) | 12.8 (1.4) | -0.46 |
| Hematocrit (%) | 37.1 (4.0) | 38.7 (3.9) | -0.40 |
| RDW, (%) | 14.0 (1.7) | 13.6 (0.8) | 0.35 |
| Neutrophils, cells/μl | 4,846.5 (1,646.6) | 5,621.5 (2,934.0) | -0.33 |
| Lymphocytes, cells/μl | 677.3 (448.4) | 926.5 (464.8) | -0.55 |
| Monocytes, cells/μl | 393.8 (295.1) | 390.3 (220.7) | 0.01 |
| Platelets, cells/μl | 206,090.9<br>(75,525.4) | 179,500.0<br>(57,479.6) | 0.40 |
| Creatinine, mg/dL | 2.6 (1.5) | 1.2 (0.3) | 1.40 |
| C-Reactive Protein, mg/L | 112.4 (46.6) | 86.4 (57.9) | 0.60 |
| D-dimer, µg/mL FEU | 1.3 (0.7) | 1.1 (0.9) | 0.23 |
| **Outcomes** |  |  |  |
| Transferred to ICU | 7 (64%) | 2 (33%) | 0.64 |
| Mechanical ventilation | 6 (55%) | 1 (17%) | 0.86 |
| Renal replacement therapy | 3 (27.3%) | 2 (33.3%) | -0.13 |
| Days in ICU | 22.1 (13.3) | 10.5 (12.0) | 0.92 |
| Hospital mortality | 6 (55%) | 1 (17%) | 0.86 |

<sup>1</sup>Continuous variables are shown as mean ± SD. Categorical variables are shown as absolute numbers (percentages). SMD: standardized mean differences.

### Global COVID-19 Perturbation of the PBMC Proteome, Plasma Proteins, and Lymphocyte Counts in Kidney Transplant Recipients

A TMT--based quantitative proteomic approach combined with liquid chromatography-tandem mass spectrometry (LC-MS/MS) was employed to quantify differentially abundant proteins (DAPs) among KTRs. The cohort was categorized based on the timing of blood sampling: patients sampled during their hospital stay (D0-wards) and those sampled approximately 30 days after discharge (CS30). These groups were compared with KTRs non-COVID-19 (KTR controls). Across seven TMT batches, a total of 2,330 proteins were identified. Focusing on proteins quantified in at least 50% of samples reduced this number to 1,403 proteins, with an average of 1,242 proteins per sample (**Supplementary Material 1**: **Supplementary Figure S1**). Among these 1,403 proteins, 363 were differentially abundant between D0-wards and KTR controls (**Supplementary Material 1**: **Supplementary Figure S2A** and **Supplementary Material 1**), while 111 showed differential abundance between CS30 and KTR controls (**Supplementary Material 1**: **Supplementary Figure S2B** and **Supplementary Material 3**).

Reactome pathway enrichment analysis revealed that D0-wards patients exhibited alterations in 15 pathways compared with KTR controls, characterized by positive normalized enrichment scores (NES) in pathways associated with interferon response, innate immune system, and neutrophil degranulation. Conversely, pathways related to glycolysis and glucose metabolism exhibited negative NES. Similar NES patterns were observed in hallmark gene set enrichment analysis, with positive NES in pathways related to interferon response and inflammatory processes, and negative NES in metabolism-related pathways (**Figure 1A**). Notably, only metabolism-related pathways remained altered in CS30 patients (**Figure 1A**).

**Figure 1.**
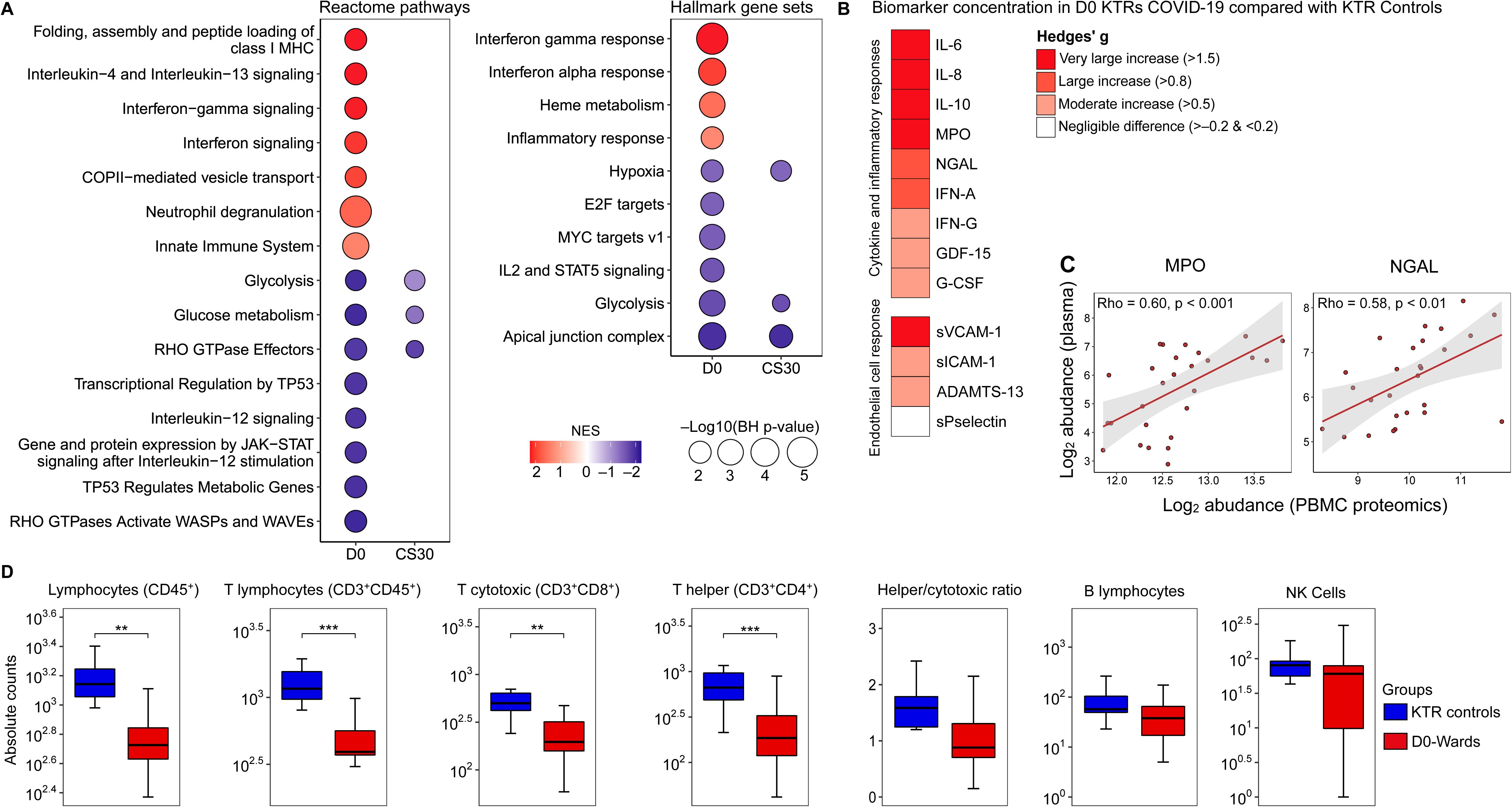
Comprehensive assessment of COVID-19 impact on PBMC proteome, plasma proteins, and lymphocyte counts. **Description Figure 1: A**) Protein set enrichment analysis (PSEA) demonstrating protein perturbations in D0-wards patients (n=17) versus KTR controls (n=10), and CS30 patients (n=6) versus KTR controls. Circle size represents the -log10(BH-adjusted p-value), while color denotes the normalized enrichment score (NES). Red indicates increased NES, and blue indicates decreased NES. **B**) Heatmap showing the magnitude of biomarker differences between D0-wards patients and KTR controls, quantified using Hedges’ g. The red gradient highlights elevated biomarker levels in D0-wards patients. (For pairwise comparisons, see Figure S3). **C**) Scatter plot illustrating Spearman’s rank correlation coefficients (Rho) between myeloperoxidase (MPO) and neutrophil gelatinase-associated lipocalin (NGAL) abundances in plasma and PBMCs (proteomic data). P-values are BH-adjusted, and **D**) Box plot comparing absolute lymphocyte counts between D0-wards COVID-19 patients and KTR controls. Statistical analyses were performed using Welch’s t-test, with significance levels indicated by **p ≤ 0.01 and ***p ≤ 0.001.

Plasma biomarker levels indicative of cytokine/inflammation and endothelial cell activation were significantly elevated in D0-wards patients (**Figure 1B**, and **Supplementary Material 1**: **Supplementary Figure S2**). Additionally, two proteins, NGAL and myeloperoxidase (MPO), were identified in both plasma and PBMCs (proteomics), demonstrating a positive moderate correlation (**Figure 1C**). Furthermore, the absolute number of lymphocytes, specifically T lymphocytes (CD3^+^CD45^+^), T cytotoxic (CD3^+^CD8^+^), and T helper (CD3^+^CD4^+^) were lower in D0-wards compared to KTR controls (**Figure 1D**). In contrast to admission samples, all plasma biomarkers (**Supplementary Material 1**: **Supplementary Table S1**) and absolute lymphocyte counts were comparable between CS30 and KTR controls (**Supplementary Material 1**: **Supplementary Table S2**), suggesting a potential return to a homeostatic state approximately 30 days after hospital discharge.

### Host response associated with acute kidney injury in KTR COVID-19 patients

We compared KTR COVID-19 patients who presented with AKI (N=6) to those who did not (N=11) during ward admission. Patients with AKI presented with higher Sequential Organ Failure Assessment (SOFA) scores (SMD: 0.90), increased C-Reactive Protein (CRP) levels (SMD: 0.60), and elevated creatinine levels (SMD: 1.40) upon ward admission. The AKI group experienced worse outcomes, including a higher rate of ICU transfers (SMD: 0.64), increased need for mechanical ventilation (SMD: 0.86), and greater hospital mortality (SMD: 0.86). Interestingly, creatinine showed a strong positive correlation with CRP (Rho=0.50, p<0.05) and SOFA respiratory subscores (Rho=0.68, p=0.002) (**Supplementary Material 1**: **Supplementary Figure S4**).

We compared the PBMC proteomic alterations in AKI and non-AKI COVID-19 patients using ssPSEA for Reactome-defined immune-related pathways. Our analysis revealed alterations in seven innate immune-related pathways, three adaptive immune system pathways, and five cytokine signaling pathways in the immune system. Additionally, no alterations were observed in hemostasis, while four metabolic pathways and one programmed cell death pathway showed alterations (**Figure 2A**). Indeed, the innate immune-related pathways exhibited the most differences when comparing patients with AKI and non-AKI; these pathways also demonstrated a pattern of positive correlation with creatinine, SOFA, and CRP levels (**Figure 2B**).

**Figure 2.**
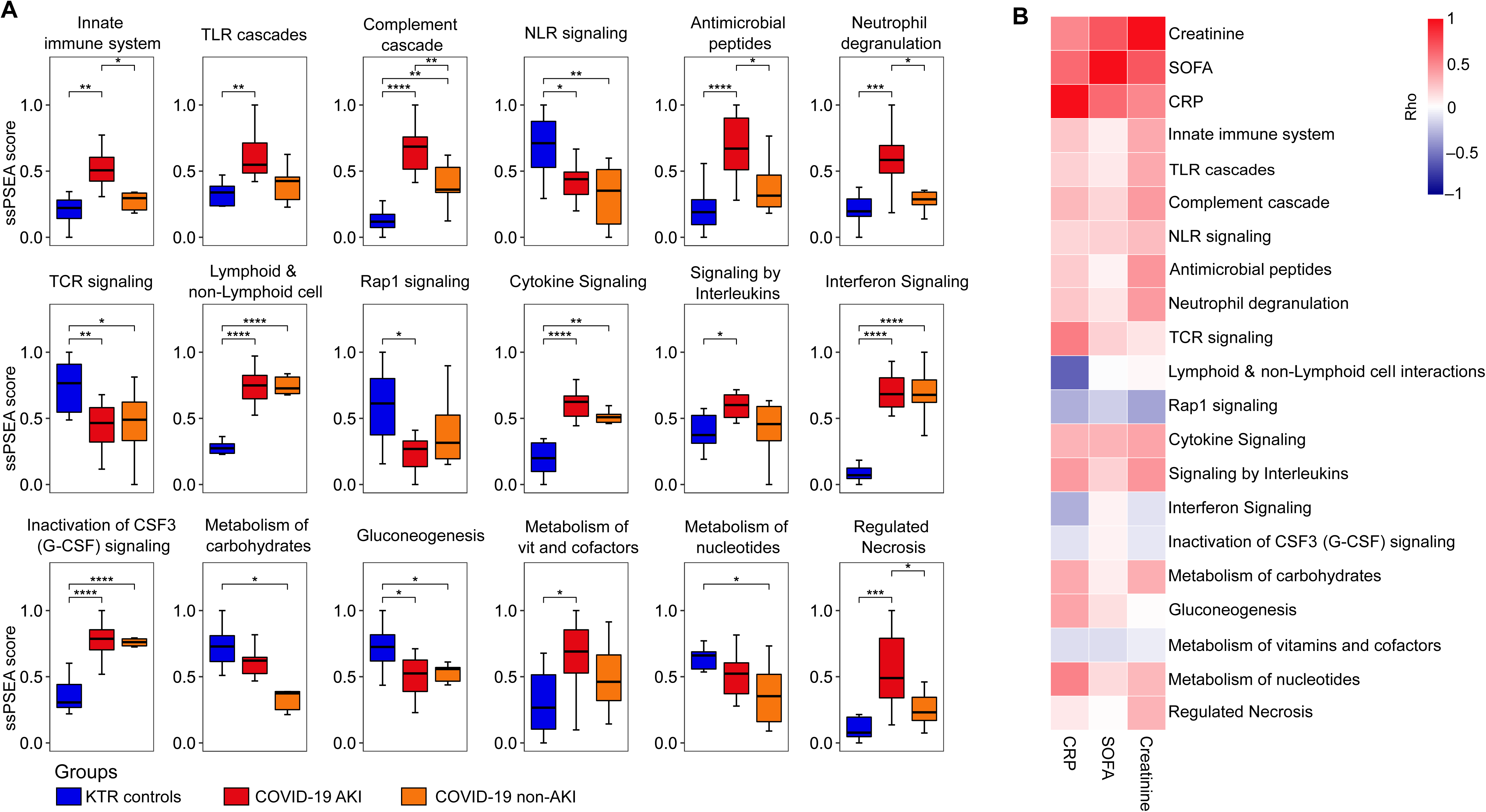
Alterations in immune-related pathways in COVID-19 patients stratified according to the presence of acute kidney injury. **Description Figure 2: A**) Single-sample protein set enrichment analysis (ssPSEA) was performed on D0-wards COVID-19 patients stratified by the presence of acute kidney injury (AKI, n=11) and non-AKI (n=6), as well as KTR controls (n=10). The figure presents only immune-related pathways that showed significant differences between the groups. Statistical analyses were conducted using ANOVA with Tukey’s post-hoc test, corrected by the Benjamini-Hochberg method. Significance is indicated as follows: *p ≤ 0.05, **p ≤ 0.01, ***p ≤ 0.001, and ****p ≤ 0.0001, and **B**) Heatmap showing Spearman’s rank correlation coefficients (Rho) between the sequential organ failure assessment (SOFA) score, C-reactive protein (CRP) levels, creatinine levels, and ssPSEA pathway scores in D0-wards COVID-19 patients (n=17). Abbreviations: TLR cascades: Toll-like receptor cascades, NLR signaling: Nucleotide-binding domain, leucine rich repeat containing receptor signaling pathway, TCR signaling: T cell receptor signaling, Immunoregulatory between Lymphoid & non-Lymphoid cell: Immunoregulatory interactions between a Lymphoid and a non-Lymphoid cell, Rap1 signaling: Ras-proximate-1 signaling and Metabolism of vit and cofactors: Metabolism of vitamins and cofactors.

Next, we explored the differences between KTR COVID-19 patients with AKI and non-AKI based on 15 host response biomarkers reflective of two key pathophysiological domains (cytokine/inflammatory response and endothelial cell activation/procoagulant responses) in plasma obtained on admission to the wards. First, we generated domain-specific PCA plots to compare the groups (**Figure 3**). There were significant differences in plasma biomarkers grouped in the cytokine/inflammatory response domain between patients with AKI and non-AKI. We also identified a substantial overlap in the endothelial cell activation/procoagulant responses between the AKI and non-AKI groups (**Figure 3A**). The complete contribution of each biomarker to a principal component (PC) score is depicted in **Supplementary Material 1**: **Supplementary Table S3**.

**Figure 3.**
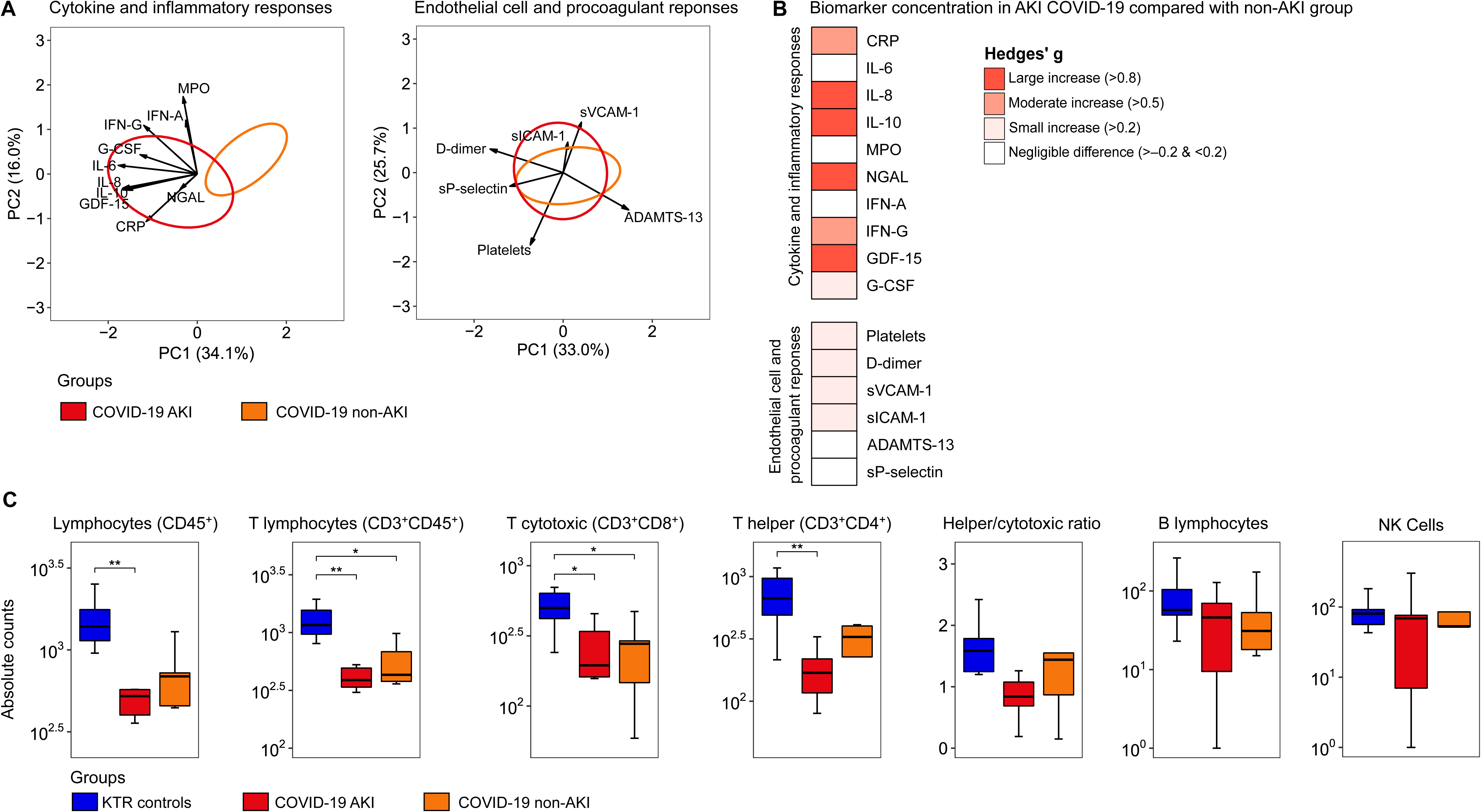
Host response biomarkers and lymphocyte count alterations in COVID-19 patients stratified according to the presence of acute kidney injury. **Description Figure 3: A)** Principal component analysis of cytokine/inflammatory responses and endothelial cell activation/procoagulant responses. The X-axis label shows the percentage of explained variance on principal component 1, while the Y-axis label shows the percentage of explained variance on principal component 2. The ellipse indicates the central 25% of the groups. The arrows indicate the direction (arrow orientation) and strength (arrow length) of the correlation between each biomarker and the PCs. The complete contribution of each biomarker to a PC score is depicted in Supplementary Table S3. **B)** Heatmap representing the Hedges’ g effect size between the AKI COVID-19 group compared with the non-AKI group (for pairwise comparison see Supplementary Table S4), and **C)** Box plot illustrating differences in lymphocyte absolute counts between D0-wards COVID-19 patients stratified according to the presence of acute kidney injury (AKI, n=11, and non-AKI, n=06) and KTR controls (n=10). Statistical analyses were performed using the ANOVA test with Tukey post-hoc test corrected by the Benjamini-Hochberg method. * Tukey post-hoc test p ≤ 0.05 and ** p ≤ 0.01. Abbreviations: IL: interleukin, GDF: growth/differentiation factor, G-CSF: granulocyte-colony stimulating factor, IFN: interferon, CRP: C-Reactive Protein, NGAL: neutrophil gelatinase-associated lipocalin, MPO: Myeloperoxidase, ADAMTS-13: ADAM metallopeptidase with thrombospondin type 1 motif 13, sP-selectin: Soluble platelet selectin, sVCAM-1: Serum vascular cell adhesion molecule-1 and sICAM-1: Soluble Intercellular Adhesion Molecule-1.

When comparing the magnitude of individual biomarker differences expressed as Hedges’ g, we observed moderate to large increases in cytokine/inflammatory markers among AKI patients, while endothelial cell activation/procoagulant markers showed small or negligible differences (**Figure 3B**). Subsequently, we evaluated the absolute number of total lymphocytes, which showed statistical differences when comparing the KTR control and COVID-19 AKI groups (**Figure 3C**). We also observed differences between the control and COVID-19 AKI groups for T helper cells. For T lymphocytes and T cytotoxic cells, differences were found in both comparisons between the KTR control vs. COVID-19 AKI groups, as well as between the KTR control vs. non-AKI groups. No differences were found in the comparison between the groups for the helper/cytotoxic ratio, B lymphocytes, and NK cells. Interestingly, for total lymphocytes, T lymphocytes, T cytotoxic cells, and T helper cells, the AKI group consistently showed lower absolute values than the non-AKI group, but with no statistical difference in the pairwise comparison.

Previous studies suggest an increase in immature cells, such as low-density neutrophils (LDNs) or granulocytic/polymorphonuclear myeloid-derived suppressor cells (PMN-MDSCs), in COVID-19 patients, which correlate with T cell suppression [24, 25]. LDNs commonly co-purify with PBMCs in various infections, enriching pathways like “neutrophil degranulation” in omics studies [26–31]. Therefore, we tested the correlation between the absolute values of T lymphocytes and the “neutrophil degranulation” ssPSEA scores, which showed a moderate negative correlation between T lymphocytes (Rho=–0.43, p=0.02), T helper cells (Rho=–0.45, p=0.03), and the “neutrophil degranulation” score (**Supplementary Material 1**: **Supplementary Figure S5**). A weak negative and not statistically significant correlation was found between T cytotoxic cells and the “neutrophil degranulation” score (Rho=–0.35, p=0.08).

## DISCUSSION

In this study, we investigated KTRs infected with SARS-CoV-2 during the initial wave of the COVID-19 pandemic in São Paulo, Brazil. Based on the reported increased severity of COVID-19 in KTRs [4, 5, 7–9], we aimed to investigate the changes in proteomics within PBMCs in this specific population. Additionally, we explored the reported immune dysregulation and endothelial cell activation/procoagulant alterations in COVID-19 [10, 32, 33] , particularly emphasizing the early stages of hospitalization while patients are in the ward. Furthermore, we investigated the reported reductions in absolute counts of lymphocytes, especially T lymphocytes and their subsets, during the acute phase of COVID-19 [34, 35] , with a specific focus on KTRs.

The proteomic response during the acute phase of COVID-19 has been widely explored [28, 30, 31, 36], while the proteomic alterations within PBMCs of KTRs were negligible. Our study revealed proteomic alterations linked to inflammatory/immune response pathways, indicated by positive NES. Interestingly, the majority of these pathways reverted to a status similar to that of non-COVID-19 KTRs after 30 days. This pattern aligns with our previous findings from a separate cohort of COVID-19 patients without KTRs [28] and is consistent with other omics reports [25, 30, 31]. Notably, the glycolysis and glucose metabolism pathways exhibited a negative NES and remained altered after 30 days. This metabolic alteration may affect immune cell function in pathogen recognition, as glycolysis plays a pivotal role in this process [37]. This dysfunctional glycolysis profile shares similarities with findings from proteomic studies of PBMCs from critically ill COVID-19 patients [28], transcriptomic/metabolic profiling of CD14+ monocytes from patients with moderate COVID-19 [38], and single-cell RNA sequencing in monocytes and macrophages from severe patients with COVID-19-associated pulmonary aspergillosis [39]. The impaired glycolysis observed in this context may serve as an indicator of the dysfunctional host response, potentially contributing to heightened morbidity and mortality rates among KTRs.

Next, we investigated the proteomic alterations based on the presence of AKI, a prevalent condition in systemic illnesses affecting a significant portion of hospitalized COVID-19 patients [40]. Analysis of immune-related pathways revealed increased NES in the innate immune system, complement cascade, regulated necrosis, antimicrobial peptides, and neutrophil degranulation in COVID-19 AKI patients. These results are in line with studies discussing the pathophysiology of COVID-19– associated AKI, which is believed to be multifaceted [40], involving factors such as local and systemic inflammatory/immune response, and activation of coagulation cascades [41, 42]. This is further evidenced when examining host response biomarkers, where the PCA corresponding to this cytokine/inflammatory response domain clearly separates patients with AKI from those without, a distinction not observed in the endothelial domain. It is noteworthy that COVID-19 pathophysiology in KTRs remains largely unknown. While various general factors commonly encountered in critically ill patients, including mechanical ventilation, hypoxia, hypotension, and exposure to nephrotoxic agents, are recognized contributors to kidney injury, [41] the role of direct viral infection with renal tropism, as proposed by some studies, remains unclear [43, 44].

Although no statistically significant differences were observed in lymphocyte counts between the AKI and non-AKI groups, it is likely that these disturbances were more severe in the AKI group, as illustrated by the finding that absolute counts of total lymphocytes and T helper cells were lower in the AKI group but not in the non-Aki group compared to the controls. Previous studies have demonstrated that T lymphocyte counts and their subsets typically decrease during the acute phase of COVID-19,[34, 35, 45]. T lymphocytes play a pivotal role in clearing the virus during SARS-CoV infection [46, 47]. Interestingly, the increase in LDNs/PMN-MDSCs may directly correlate with the decrease in the number of T cells [24, 25]. Despite their immunosuppressive properties, LDNs/PMN-MDSCs have also been associated with a pro-inflammatory phenotype, characterized by elevated levels of cytokine secretion and formation of neutrophil extracellular traps [27, 48, 49]. The presence of LDNs has been observed in previous studies [26, 28, 29], , and might be correlated to the severity of infectious diseases [25, 30]. In fact, the AKI group is more severe than the non-AKI, showing higher SOFA scores, CRP levels, and worse outcomes. This may be associated with high scores for LDNs, as well as inflammatory/immune responses. Our findings suggest a relationship between T-cell suppression and elevated levels of LDN-related proteins in PBMCs during COVID-19, particularly in patients with AKI. This association may carry important therapeutic implications for managing immune dysregulation in KTRs, highlighting the need for targeted interventions aimed at mitigating these immune alterations.

This study has some limitations. First, the sample size was relatively small, and all patients were enrolled at a single center. Additionally, the absence of a validation cohort from another hospital in a different geographical region limits the generalizability of our findings. Moreover, we did not investigate the influence of viral load or the roles of virus variants on the results obtained. Our study was conducted in 2020, so the relevance to current variants is less clear, since SARS-CoV2 variants exhibit increasing fitness, and declining pathogenicity and induced-inflammatory response [50]. Furthermore, functional assays to validate glycolysis metabolism changes and/or the potential contribution of LDNs in AKI were not conducted, which could have provided further insight into our results. Despite these limitations, our study also has notable strengths. We investigated PBMCs, a cell type that is not often explored in COVID-19 proteomics research. Our study of KTRs infected with SARS-CoV-2, along with a well-established cohort of controls, provides valuable insights and is relatively unexplored. Additionally, all individuals were unvaccinated, allowing for a clearer assessment of proteomic alterations in the absence of vaccination. Furthermore, the samples obtained in the wards and after discharge provided a comprehensive understanding of cellular proteomic changes throughout the initial symptoms of the disease and convalescence.

Future studies with larger, more diverse cohorts are needed to validate our findings and evaluate their long-term clinical implications. As a next step, we aim to explore patients with community-acquired infections, including both KTRs and non KTRs, to investigate broader mechanisms of immune dysregulation. These insights lay the groundwork for the development of targeted therapeutic interventions to address immune dysregulation in vulnerable populations, particularly kidney transplant recipients.

In summary, our study offers preliminary insights into the proteomic alterations within PBMCs of KTRs infected with SARS-CoV-2 during the early phase of the COVID-19 pandemic. We identified significant dysregulation in key biological pathways, particularly those involved in glycolysis, glucose metabolism, and neutrophil degranulation, which may reflect the heightened inflammatory state associated with COVID-19 in this vulnerable population. While our findings suggest an altered immune response—characterized by elevated cytokines, inflammatory mediators, and decreased lymphocyte counts—these results must be interpreted cautiously due to the small sample size. The observed association between T-cell suppression and increased levels of LDN-related proteins in AKI patients warrants further investigation, as it may hold therapeutic implications for mitigating immune dysregulation in COVID-19-infected KTRs.

## Data Availability Statement

Raw and processed mass spectrometry proteomic data generated in this study are available at ProteomeXchange with the dataset identifier PXD051702.

## Author Contributions

Giuseppe G. F. Leite (Conceptualization; Proteomic sample preparation; Data curation; Formal analysis; Investigation; Methodology; Visualization; Writing original draft; Writing – review & editing);

Mônica Bragança Sousa (Involved in collecting the data; Writing – review & editing);

Larissa de Oliveira C. P. Rodrigues (Involved in collecting the data; Writing – review & editing);

Milena Karina Colo Brunialti (Involved in collecting the data; Writing – review & editing); José Medina-Pestana Writing (Intellectual input; Writing – review & editing);

Joe M. Butler Writing (Methodology; Intellectual input; Writing – review & editing); Hessel Peters-Sengers (Methodology; Intellectual input; Writing – review & editing);

Lúcio Requião-Moura (Supervision - Intellectual input; Data curation; Writing original draft; Writing – review & editing);

Reinaldo Salomão (Funding acquisition; Supervision - Intellectual input; Data curation; Visualization; Writing original draft; Writing – review & editing)

## Acknowledgments

The authors thank Nancy Bellei, Universidade Federal de São Paulo, for the SARS-CoV-2 PCR test; Paulo R. Abrão Ferreira, Jaquelina Sonoe Ota-Arakaki and Paula M. Peçanha-Pietrobom for the support with the selection and enrollment of patients. We thank Paolo Di Mascio and Graziella E. Ronsein for the mass spectrometry analyses performed at the Redox Proteomics Core of the Mass Spectrometry Resource at Chemistry Institute, University of São Paulo (FAPESP grant numbers 2012/12663-1 and 2016/00696-3, CEPID Redoxoma 2013/07937-8).

