## Supplementary Material 1 for "Proteomic profiling of peripheral blood mononuclear cells reveals immune dysregulation and metabolic alterations in kidney transplant recipients with COVID-19"

### MEASUREMENTS

#### Plasma biomarker measurements

Biomarkers indicative of host response domains implicated in COVID-19 pathogenesis were assessed upon admission to the wards and 30 days post-hospital discharge. Utilizing the Cytometric Bead Array (CBA) Flex Set kits, Interleukin (IL)-6, IL-8, IL-10, G-CSF (granulocyte colony-stimulating factor), interferons-alpha (IFN-α), and -gamma (IFN-γ) were quantified. The analysis of these samples was conducted on the LSRFortessa instrument from BD Biosciences, San Jose, CA, USA. Disintegrin and metalloproteinase with thrombospondin motifs 13 (ADAMTS-13), growth/differentiation factor 15 (GDF-15), soluble intercellular adhesion molecule-1 (sICAM-1), myeloperoxidase (MPO), P-selectin, neutrophil gelatinase-associated lipocalin (NGAL), soluble circulating vascular cell adhesion molecule-1 (sVCAM-1) were measured in plasma by flow cytometry (MAGPIX® Instrument, Luminex Corporation, Austin, TX, USA), using the kit Milliplex Map Human Cardiovascular Disease (CVD) Magnetic Bead Panel 2 - Cardiovascular Disease Multiplex Assay (Temecula, CA, USA). Additionally, D-dimer and C-reactive protein (CRP) levels were determined through immunoturbidimetric assay (Roche Diagnostics). To complement this investigation, we assessed the plasma levels of the mentioned biomarkers in 10 age- and gender-matched kidney transplant recipients (KTRs) volunteers, except for CRP and D-dimer.

**Absolute Counts of Leukocytes in Peripheral Blood**

NK cells, B, T lymphocytes, and TCD4+ and TCD8+ subpopulations cell counts. Absolute cell numbers of lymphocytes were obtained by Flow Cytometry using Multitest reagents and Trucount Absolute Counting Tubes (BD Biosciences, San Jose, CA, USA) according to the manufacturer. One tube was used for B and NK cells (BD MultiTEST™ CD3 fluorescein isothiocyanate (FITC)/CD16+CD56 phycoerythrin (PE)/CD45 peridinin chlorophyll protein (PerCP)/CD19 allophycocyanin (APC) and another for T lymphocytes subpopulations (BD Multitest™ CD3 fluorescein isothiocyanate (FITC)/CD8 phycoerythrin (PE)/CD45 peridinin chlorophyll protein (PerCP)/CD4 allophycocyanin (APC). Briefly, aliquots of 50uL of EDTA-treated blood were added to each tube and incubated in the dark at room temperature for 15 min, followed by erythrocyte lyses with 450 µL of 1X BD FACS lysing solution. Samples were run in a FACSCalibur flow cytometer and analyzed with BD Multiset Software.

**Proteomics**

*Preparation of PBMCs and protein extraction and digestion*

Protein samples were prepared as previously described [1] with minor changes. Briefly, PBMCs were thawed, and the protein extracts were obtained by lysis in 7 M urea, 2 M thiourea, and 200 mM Dithiothreitol (DTT, Sigma Aldrich, St. Louis, MO, USA) with the Protease Inhibitor Mix (Cytiva, Marlborough, MA, USA). After centrifugation at 16, 000 g for 10 min at 4°C, the protein concentration in the supernatants was determined using the Bradford method [2]. The samples were reduced with 5 mM DTT at 65°C for 30 min and then alkylated with 15 mM iodoacetamide (Sigma Aldrich, St. Louis, MO, USA) at room temperature for 30 min in the dark. The proteins were precipitated in acetone: methanol (8:1, v:v) at −80°C overnight (16 h) and, after two washes with methanol, recovered by centrifugation at 14, 000 g for 10 min at 4°C. They were then dissolved in 100 mM triethylammonium bicarbonate buffer (TEAB, Thermo Scientific, Waltham, MA, USA) to a protein concentration of 1 μg.μL$^{-1}$. Trypsin/Lys-C Mix (Promega, Madison, WI, USA) was added at a 1:50 enzyme:protein ratio at 37°C, and samples were digested overnight (16 h). The peptides were then desalted using PolyLC C18 tips (PolyLC Inc., Waltham, MA, USA), vacuum-dried, and stored at −80°C.

*TMT labeling*

After protein extraction and digestion, the peptides were dissolved in 100 mM of TEAB buffer. The peptide concentrations were measured using Quantitative Colorimetric Peptide Assay (Thermo Scientific, Waltham, MA, USA) before TMT labeling to ensure that the amounts of peptides in each channel for TMT labeling were equal. TMT labeling was performed according to the manufacturer's recommendations with minor modifications. For tag reconstitution, the TMT reagent (TMT sixplex™ Isobaric Label Reagent Set, Thermo Scientific, Waltham, MA, USA) was dissolved in 41 μl acetonitrile (Sigma Aldrich, St. Louis, MO, USA) according to the manufacturer’s instructions. Each batch of the TMT experiment contained five different samples and one global internal standard (GIS) sample created by pooling PBMC samples from all individuals of the cohort (TMT channel 126 was used to label GIS). From every sample, 25 μg was labeled with 10 μl of a TMT tag. Reactions were incubated at room temperature for 1 h. The labeling reaction was quenched by an additional 4 μl of 5% hydroxylamine for 15 min. The TMT-labeled samples were pooled with a protein concentration ratio of 1:1:1:1:1:1 [3]. Each mixture was dried and stored at −20°C until the liquid chromatography coupled to tandem mass spectrometry (LC-MS/MS) analysis.

*LC-MS/MS analysis*

Samples were analyzed by LC-MS/MS using an Orbitrap Fusion Lumos mass spectrometer (Thermo Scientific, Waltham, MA, USA) coupled to a Nano EASY-nLC 1200 (Thermo Scientific, Waltham, MA, USA). TMT-labeled peptides were injected into a trap column (nanoViper C18, 3 μm, 75 μm × 2 cm, Thermo Scientific, Waltham, MA, USA) with 12 μL of solvent A (0.1% formic acid) at 980 bar. The trapped peptides were eluted onto a C18 column (nanoViper C18, 2 μm, 75 μm × 15 cm) at a flow rate of 300 nL/min and subsequently separated with a 5%–28% acetonitrile gradient with 0.1% formic acid for 80 min, followed by a 28%–40% acetonitrile gradient with 0.1% formic acid for 10 min. The electrospray ionization source was operated in a positive mode, with voltage and temperature being adjusted to 2.1 kV and 300°C, respectively. The mass spectrometer was operated in a data-dependent acquisition mode, with the MS scan in the m/z range of 400–1600 (with a target value of $10^6$ ions) using the Orbitrap analyzer at a resolution of 120, 000 (at m/z 400), followed by higher-energy collisional dissociation (set to 38%) of the 10 most intense ions at a resolution of 50, 000. The isolation window for precursor ions was set to 0.7 m/z, the minimum count to trigger MS/MS events was 25, 000 counts per second, and the dynamic exclusion time was set to 60 s.

*Proteomic data processing*

Raw data files from Orbitrap Fusion Lumos were processed using Proteome Discoverer Suite version 3.0 (Thermo Scientific, Waltham, MA, USA, formic acid). Peptides were identified using the SEQUEST HT search engine with the UniProtKB/Swiss-Prot (TaxID = 9606, *Homo sapiens*, 20, 523 sequences) database and a list with common contaminants (245 sequences). The following search settings were applied: precursor mass tolerance of 10 ppm, fragment ion tolerance of 0.02 Da (MS2 mode), fully tryptic specificity, maximum of two missed cleavages, minimum peptide length of 6 and maximum peptide length of 144, TMT-labeled peptide *N*-terminals and lysines (+229.163 Da), carbamidomethylation of cysteine (+57.021 Da) as a fixed modification, and oxidation of methionine residues (+15.994 Da) as a variable modification. The false discovery rate for proteins, peptides, and peptide spectral matches was set to 1%. TMT reporter ions were matched with a 20 ppm tolerance window, and both unique and razor peptides were considered for quantitation. The abundance was normalized on “Total Peptide Amount” and then scaled with “On Controls Average” (TMT channel 126 was used as a reference, GIS) [4]. In this case, it summed the peptide group abundance for each sample, determined the maximum sum for all files, and calculated the normalization factor using the sum of the sample and the maximum sum in all files [5].

*Batch effect correction*

The matrix of Proteome Discoverer normalized abundance of TMT reporters was imported into the R software environment (version 4.2.0) and was log2 transformed. Proteins quantified in ≥ 50% of samples were included in subsequent analyses. The filtered data were then imputed by applying the function “rfImpute” present in “randomForest” R packages (version 4.7-1.1), as previously reported [6,7]. Subsequently, ComBat (R sva package, version) was used to remove variability due to multiple batches [8].

### Supplementary Tables

#### Supplementary Table S1. Pairwise comparison of plasma biomarkers in convalescent sample 30 days post-hospital discharge

| | **CS30** (N=6) | **KTR Controls** (N=10) | ***p*-value** |
|---|---|---|---|
| **Cytokine and inflammatory responses** | | | |
| IL-6, pg/mL (mean, SD) | 7.2 (6.5) | 6.2 (3.7) | 0.75 |
| IL-8, pg/mL (mean, SD) | 6.5 (2.3) | 8.8 (5.5) | 0.26 |
| IL-10, pg/mL (mean, SD) | 2.8 (2.4) | 2.1 (1.8) | 0.59 |
| MPO, ng/mL (mean, SD) | 26.7 (12.4) | 15.3 (6.5) | 0.07 |
| NGAL, ng/mL (median, IQR) | 62.1 (44.1, 84.3) | 52.3 (38.2, 88.9) | 0.86 |
| IFN-A, pg/mL (mean, SD) | 0.6 (1.2) | 1.9 (3.1) | 0.38 |
| IFN-G, pg/mL (median, IQR) | 1.0 (0.2, 1.2) | 1.4 (0.5, 2.1) | 0.62 |
| GDF-15, ng/mL (median, IQR) | 1.3 (1.3, 1.7) | 1.4 (1.0, 3.0) | 0.7 |
| G-CSF, pg/mL (mean, SD) | 13.5 (2.6) | 12.2 (4.4) | 0.47 |
| **Endothelial cell and procoagulant responses** | | | |
| sVCAM-1, ng/mL (mean, SD) | 838.1 (332.8) | 787.4 (223.3) | 0.79 |
| sICAM-1, ng/mL (mean, SD) | 64.0 (38.2) | 76.7 (59.7) | 0.61 |
| ADAMTS-13, ng/mL (median, IQR) | 704.8 (550.0, 803.1) | 551.9 (513.5, 620.4) | 0.42 |
| sP-selectin, ng/mL (mean, SD) | 75.1 (46.3, 105.7) | 68.6 (51.7, 84.7) | 0.92 |

#### Supplementary Table S2. Lymphocyte counts in convalescent sample 30 days post-hospital discharge

| | **CS30** (N=6) | **KTR Controls** (N=10) | ***p*-value** |
|---|---|---|---|
| Lymphocytes (CD45+) | 1,752.5 (830.1) | 1,510.6 (486.7) | 0.53 |
| T lymphocytes (CD3+CD45+) | 1,493.5 (830.9) | 1,265.2 (386.2) | 0.55 |
| T cytotoxic (CD3+CD8+) | 685.8 (400.5) | 511.7 (150.9) | 0.34 |
| T helper (CD3+CD4+) | 649.3 (470.7) | 687.7 (339.9) | 0.86 |
| Helper/cytotoxic ratio | 1.3 (0.8) | 1.5 (0.7) | 0.63 |
| B lymphocytes | 84.3 (46.4) | 121.9 (146.2) | 0.46 |
| NK Cells | 137.2 (76.7) | 92.2 (49.8) | 0.23 |

Continuous variables are shown as mean ± SD.

**Supplementary Table S3. Contribution of each biomarker to principal components 1 and 2 in each host response domain in patients admitted to wards with COVID-19 stratified according to the presence of acute kidney injury**

| Cytokine and inflammatory responses | | |
|---|---|---|
| **Biomarker** | **PC1** | **PC2** |
| IL-6 | 15.68 | 2.71 |
| IL-10 | 14.77 | 4.47 |
| GDF-15 | 14.70 | 5.06 |
| IL-8 | 14.15 | 5.09 |
| G-CSF | 11.36 | 6.08 |
| IFN-G | 10.62 | 15.33 |
| CRP | 10.16 | 15.01 |
| NGAL | 3.46 | 4.89 |
| MPO | 2.81 | 24.39 |
| IFN-A | 2.30 | 16.97 |
| **Endothelial cell and procoagulant responses** | | |
| **Biomarker** | **PC1** | **PC2** |
| D-dimer | 29.50 | 10.36 |
| ADAMTS-13 | 26.46 | 16.35 |
| sP-selectin | 21.53 | 5.96 |
| Platelets | 13.28 | 31.77 |
| sVCAM-1 | 7.37 | 22.17 |
| sICAM-1 | 1.85 | 13.39 |

IL: interleukin, GDF: growth/differentiation factor, G-CSF: granulocyte-colony stimulating factor, IFN: interferon, CRP: C-Reactive Protein, NGAL: neutrophil gelatinase-associated lipocalin, MPO: Myeloperoxidase, ADAMTS-13: ADAM metallopeptidase with thrombospondin type 1 motif 13, sP-selectin: Soluble platelet selectin, sVCAM-1: Serum vascular cell adhesion molecule-1 and sICAM-1: Soluble Intercellular Adhesion Molecule-1.

**Supplementary Table S4. Pairwise comparison of plasma biomarkers in patients admitted to wards with COVID-19 stratified according to the presence of acute kidney injury**

| Characteristic | **AKI** (N=11)[1] | **non-AKI** (N=6)[1] | ***p*-value** |
|---|---|---|---|
| **Cytokine and inflammatory responses** | | | |
| C-Reactive Protein, mg/L | 112.4 (46.6) | 86.4 (57.9) | 0.30 |
| IL-6, pg/mL (mean, SD) | 185.4 (211.4) | 109.2 (105.1) | 0.59 |
| IL-8, pg/mL (mean, SD) | 36.3 (12.8) | 22.8 (15.7) | 0.1 |
| IL-10, pg/mL (mean, SD) | 11.1 (5.9) | 6.7 (3.8) | 0.07 |
| MPO, ng/mL (mean, SD) | 97.5 (45.2) | 90.5 (24.6) | 0.73 |
| NGAL, ng/mL (median, IQR) | 58.0 (50.2, 90.5) | 160.3 (94.9, 189.0) | **0.02** |
| IFN-A, pg/mL (mean, SD) | 13.1 (15.2) | 21.8 (34.4) | 0.57 |
| IFN-G, pg/mL (median, IQR) | 4.0 (2.4, 9.9) | 1.8 (0.3, 6.5) | 0.29 |
| GDF-15, ng/mL (median, IQR) | 4.2 (2.4, 10.0) | 2.0 (1.5, 2.2) | **0.01** |
| G-CSF, pg/mL (mean, SD) | 21.7 (11.8) | 17.9 (8.6) | 0.45 |
| **Endothelial cell and procoagulant responses** | | | |
| Platelets, cells/µl (mean, SD) | 206,090.9 (75,525.4) | 179,500.0 (57,479.6) | 0.43 |
| D-dimer, µg/mL FEU (mean, SD) | 1.3 (0.7) | 1.1 (0.9) | 0.44 |
| sVCAM-1, ng/mL (mean, SD) | 3,591.7 (2,854.6) | 2,856.9 (2,016.9) | 0.54 |
| sICAM-1, ng/mL (mean, SD) | 184.1 (152.3) | 97.7 (48.8) | 0.1 |
| ADAMTS-13, ng/mL (median, IQR) | 715.3 (182.0) | 685.1 (172.7) | 0.66 |
| sP-selectin, ng/mL (mean, SD) | 82.7 (36.2) | 78.8 (24.6) | 0.79 |

### Supplementary Figures

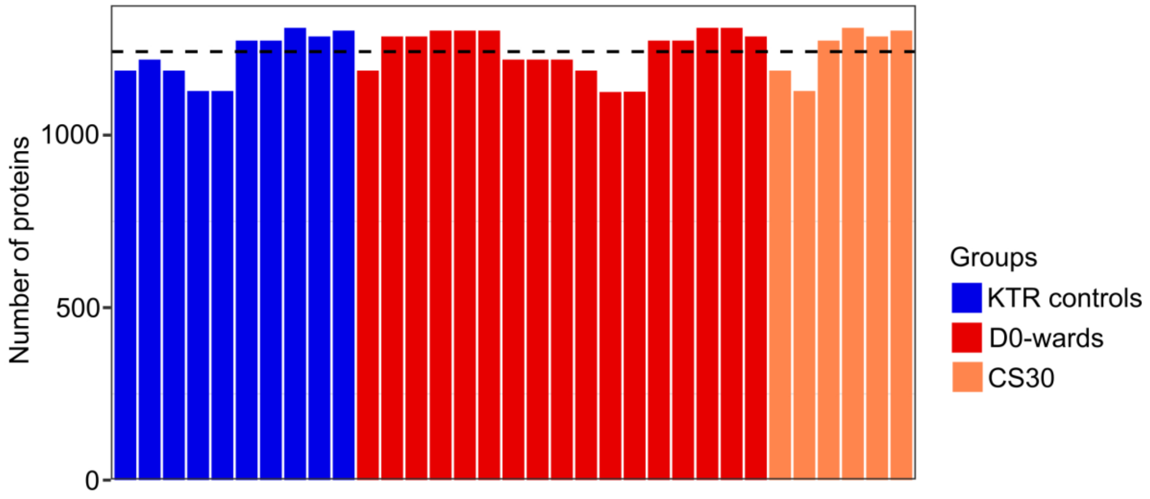


**Supplementary Figure S1. Overview of quantitative proteomics.**

**Description Figure S1:** Barplot showing proteins identified in each sample; the dotted line represents the average number of proteins per sample (1,242).

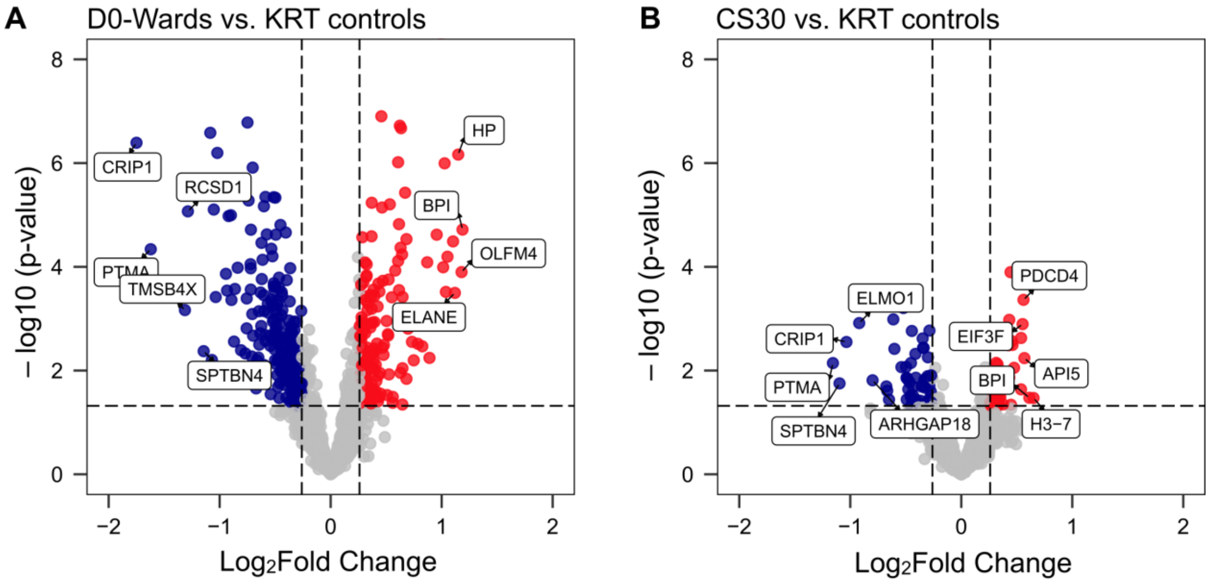


**Supplementary Figure S2. Volcano plots of differential protein abundance in kidney transplant recipients (KTRs) compared with KTRs without COVID-19**

**Description Figure S2: A**) Volcano plots showcase variations in PBMCs proteomic responses between D0-Wards KRT patients and KTR controls and **B**) Convalescent sample approximately 30 after discharge (CS30) vs. KTR controls. Proteins are identified as differentially abundant with a p-value < 0.05 and log2 fold change < −0.26 or > 0.26. The protein names highlight the top five with high abundance (in red) and the top five with low abundance (in blue) genes. Gray dots represent genes that did not show differential expression between groups.

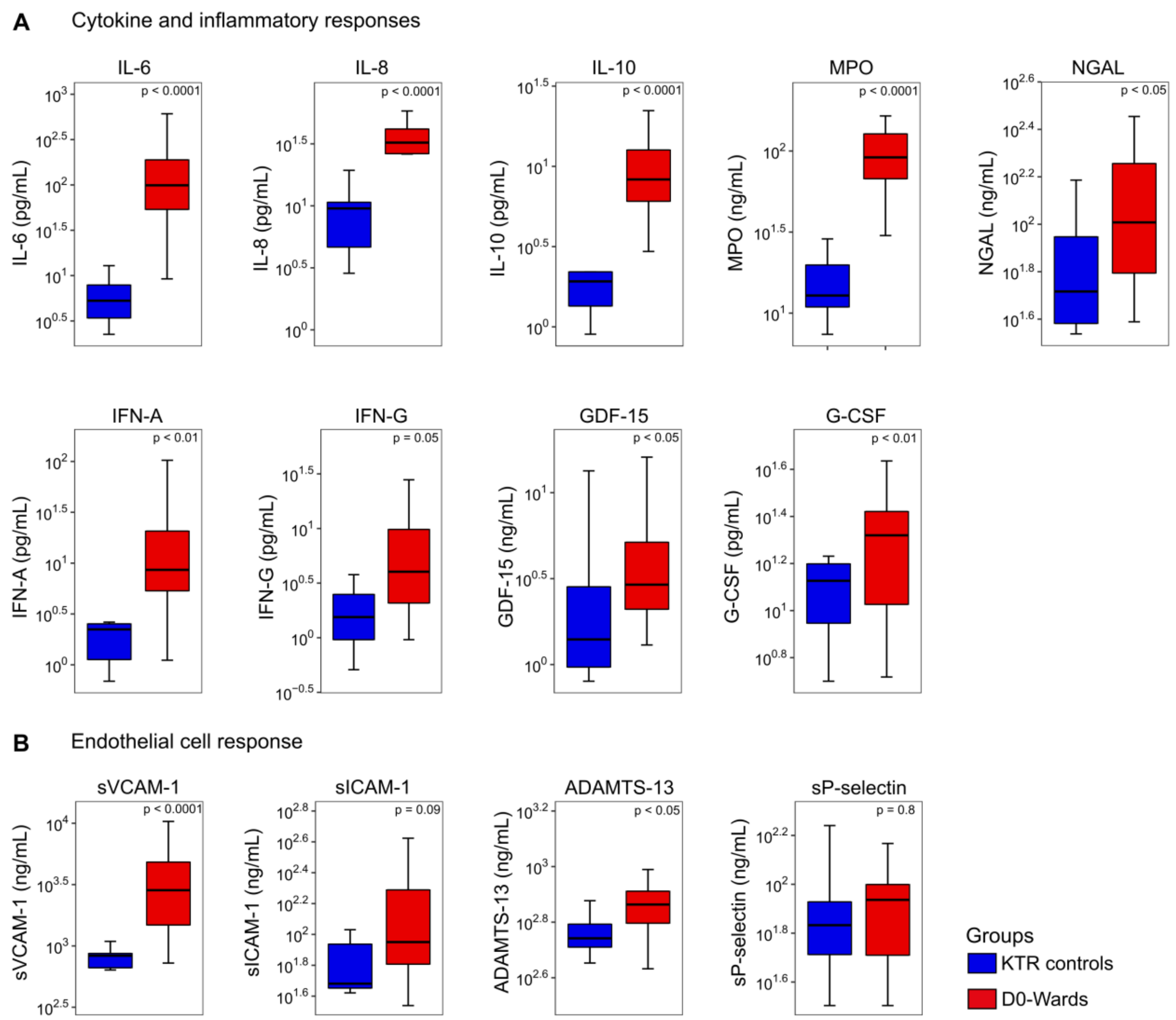


**Supplementary Figure S3. Host response biomarkers of kidney transplant recipient patients stratified by SARS-CoV-2 infection status.**

**Description figure S3: A)** Alterations in cytokine/inflammatory response-related biomarkers in KTR COVID-19-positive patients admitted to wards (D0-Wards) and KTR controls, **B)** Alterations in endothelial cell response-related biomarkers. Statistical analyses were performed using Welch's t-test or Mann-Whitney U test.

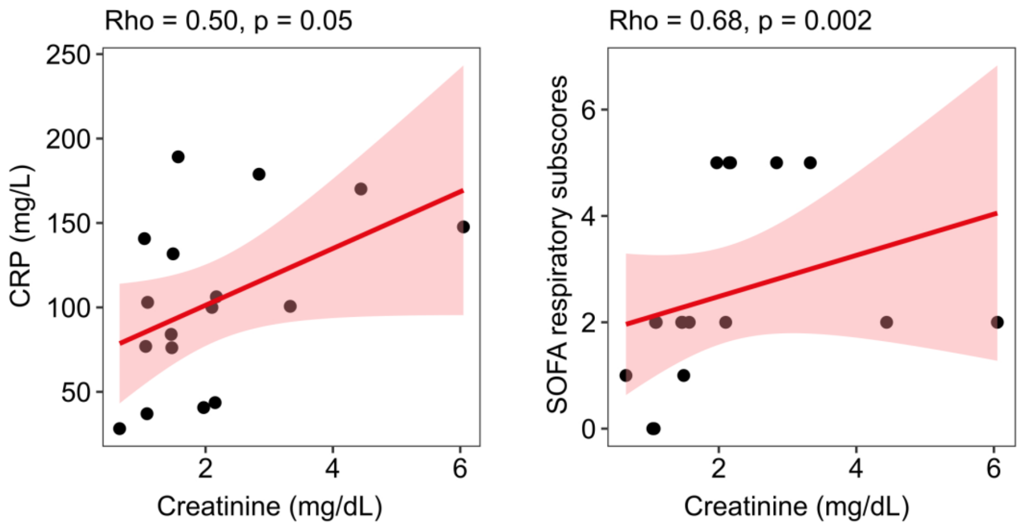


**Supplementary Figure S4**. Correlation between creatinine, CRP, and SOFA score.

**Description figure S4:** Scatter plots showing the Spearman's rank correlation coefficients (Rho) between C-Reactive Protein (CRP) levels, the sequential organ failure assessment score (SOFA score), and creatinine levels in D0-wards patients (n=17).

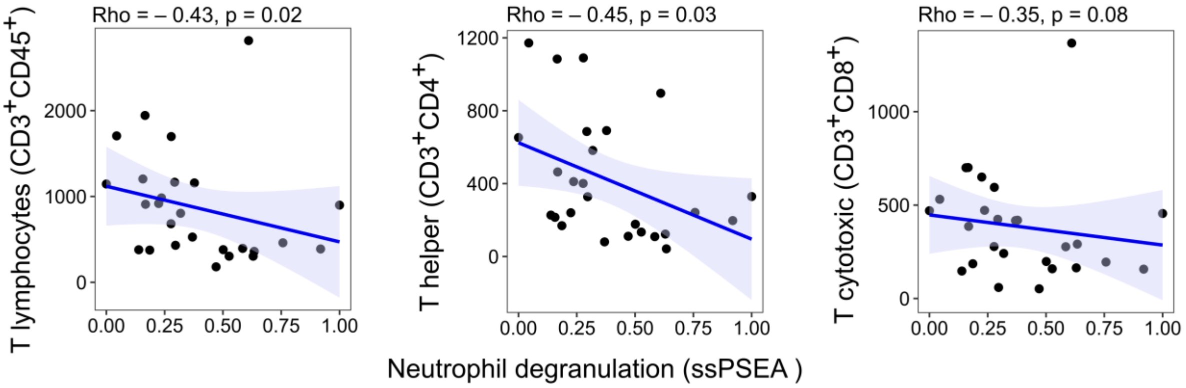


**Supplementary Figure S5.** Correlation between the absolute values of T lymphocytes, T helper, T cytotoxic, and "neutrophil degranulation" ssPSEA scores.

**Description figure S5:** Scatter plots showing the Spearman's rank correlation coefficients (Rho) between tween the absolute values of T lymphocytes, T helper, T cytotoxic and "neutrophil degranulation" ssPSEA scores in D0-wards patients (n=17) and KTR controls (n=10)

8. Gutierrez-Quiceno L, Dammer EB, Johnson AG, et al. A proteomic network approach resolves stage-specific molecular phenotypes in chronic traumatic

encephalopathy. *Molecular Neurodegeneration*. 2021;16(1):40. doi:10.1186/s13024-021-00462-3
